# Brain eQTLs of European, African American, and Asian ancestry improve interpretation of schizophrenia GWAS

**DOI:** 10.1101/2024.02.13.24301833

**Authors:** Yu Chen, Sihan Liu, Zongyao Ren, Feiran Wang, Yi Jiang, Rujia Dai, Fangyuan Duan, Cong Han, Zhilin Ning, Yan Xia, Miao Li, Kai Yuan, Wenying Qiu, Xiao-Xin Yan, Jiapei Dai, Richard F. Kopp, Jufang Huang, Shuhua Xu, Beisha Tang, Eric R. Gamazon, Tim Bigdeli, Elliot Gershon, Hailiang Huang, Chao Ma, Chunyu Liu, Chao Chen

## Abstract

Research on brain expression quantitative trait loci (eQTLs) has illuminated the genetic underpinnings of schizophrenia (SCZ). Yet, the majority of these studies have been centered on European populations, leading to a constrained understanding of population diversities and disease risks. To address this gap, we examined genotype and RNA-seq data from African Americans (AA, n=158), Europeans (EUR, n=408), and East Asians (EAS, n=217). When comparing eQTLs between EUR and non-EUR populations, we observed concordant patterns of genetic regulatory effect, particularly in terms of the effect sizes of the eQTLs. However, 343,737 cis-eQTLs (representing ∼17% of all eQTLs pairs) linked to 1,276 genes (about 10% of all eGenes) and 198,769 SNPs (approximately 16% of all eSNPs) were identified only in the non-EUR populations. Over 90% of observed population differences in eQTLs could be traced back to differences in allele frequency. Furthermore, 35% of these eQTLs were notably rare (MAF < 0.05) in the EUR population. Integrating brain eQTLs with SCZ signals from diverse populations, we observed a higher disease heritability enrichment of brain eQTLs in matched populations compared to mismatched ones. Prioritization analysis identified seven new risk genes (*SFXN2*, *RP11-282018.3*, *CYP17A1*, *VPS37B*, *DENR*, *FTCDNL1*, and *NT5DC2*), and three potential novel regulatory variants in known risk genes (*CNNM2*, *C12orf65*, and *MPHOSPH9*) that were missed in the EUR dataset. Our findings underscore that increasing genetic ancestral diversity is more efficient for power improvement than merely increasing the sample size within single-ancestry eQTLs datasets. Such a strategy will not only improve our understanding of the biological underpinnings of population structures but also pave the way for the identification of novel risk genes in SCZ.

## Introduction

Genome-wide association studies (GWAS) have identified 287 risk loci associated with schizophrenia (SCZ)^1^. Yet, the underlying mechanisms of these loci in disease development and progression remain poorly understood. Primarily, over 80% of GWAS risk loci reside in non-coding regions, devoid of protein-coding sequences, making it challenging to attribute them to specific genes. Moreover, predicting the regulatory effect of these loci proves challenging due to their tendency for gene-specific and tissue-specific effects. One effective strategy for gaining insights into their functions involves the integration of SCZ GWAS signals with expression quantitative trait loci (eQTLs), utilizing genotype and expression data from post-mortem brains. These brain eQTLs establish crucial links between risk genomic regions and gene expression levels, prioritizing potential disease risk genes through methods such as colocalization and transcriptome-wide association study (TWAS).

Past brain eQTL studies primarily focused on European (EUR) ancestry^2–6^. Global population diversity has not been adequately represented. Cross-population studies have shown that these European ancestry based models do not effectively predict gene expression in other ancestral groups^7^. This limitation weakens the power to detect TWAS associations in genetically diverse samples. While multi-ancestry eQTL meta-analyses in the human brain improve statistical power in uncovering risk loci shared across populations, key genetic variants regulating expression in specific underrepresented populations remain largely uncharted. The benefits of having brain eQTLs in diverse populations have not been thoroughly documented. Identifying eQTLs specific to biomedically underrepresented groups like African Americans (AA) and East Asians (EAS) can better understand the genetic contributions to disease susceptibilities and outcomes in these populations^7^. These populations have unique genetic variants and linkage disequilibrium (LD) patterns. Additionally, previous studies have shown that combining eQTLs from different ancestries can enable fine-mapping of causal variants and uncover potential novel mechanisms of brain disorders^8,9^. Thus, the question of how to effectively leverage difference to uncover potential novel mechanisms of brain disorders is a significant topic in the field.

To enhance the diversity in brain eQTL mapping and improve the interpretation of SCZ GWAS across populations, we performed brain eQTL mapping in three major ancestries. Our data pool comprised genotype and RNA-seq data of AA (n= 158) and EUR (n= 408) from the PsychENCODE Consortium and EAS (n= 217) from the Chinese Human Brain Bank (Supplementary Table 1). We juxtaposed non-EUR results against EUR to systematically examine differences and similarities in the brain eQTLs. Further, we investigated the contributing factors for eQTL differences across populations. By applying diverse population brain eQTLs to TWAS and colocalization analysis of SCZ GWAS, we identified new risk genes and pathways. Lastly, we identified likely causal variants by multi-ancestry fine-mapping. The two key questions we sought to answer are: 1) what drives the brain eQTL differences across populations? 2) what do we gain by studying brain eQTLs in diverse populations?

## Results

To capture brain eQTLs across diverse populations, we utilized high-density genotype data alongside high-throughput RNA-sequencing from prefrontal cortices. We derived AA (n = 158) and EUR (n = 408) data from the BrainGVEx project of the PsychENCODE Consortium. We generated EAS (n = 217) data from the Chinese Human Brain Bank (Fig. 1). Following rigorous quality checks and preprocessing (Extended Data Fig. 1 and Extended Data Fig. 2), we compiled expression data for 18,939 genes and genotype data at 6.4 million autosomal single nucleotide polymorphisms (SNPs) across the three groups. Aligning the samples with the 1000 Genomes Project reference populations, principal component analysis (PCA) confirmed the ancestry origins of donors (Fig. 2a). We ensured sample identity consistency by comparing the genotypes from the DNA and RNA samples. See methods for additional details.

**Fig. 1.**
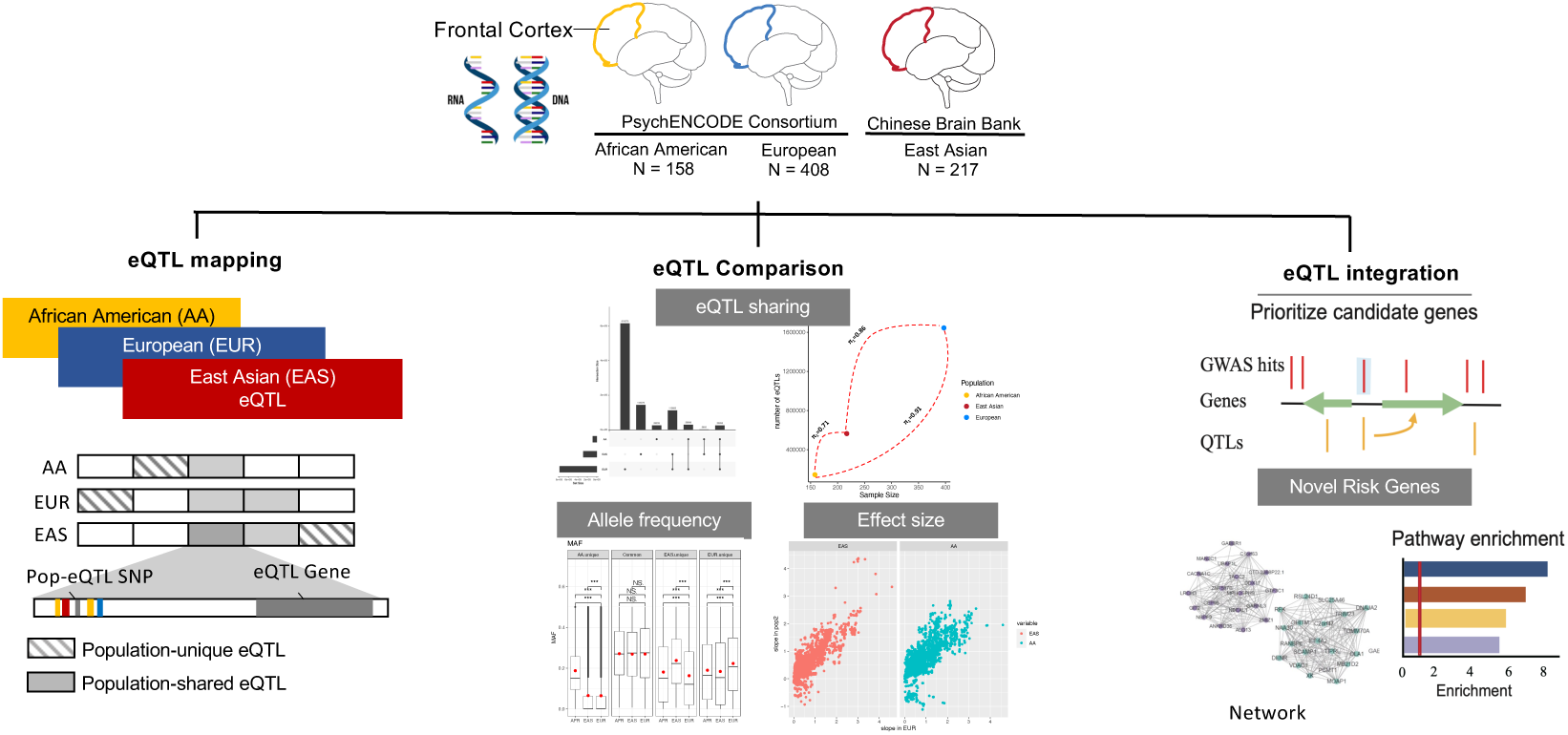
Study design. We examined genotype and RNA-seq data from African Americans (AA), Europeans (EUR), and East Asians (EAS) to identify expression quantitative trait loci (eQTLs) unique to non-European populations and their role in schizophrenia risk.

**Fig. 2:**
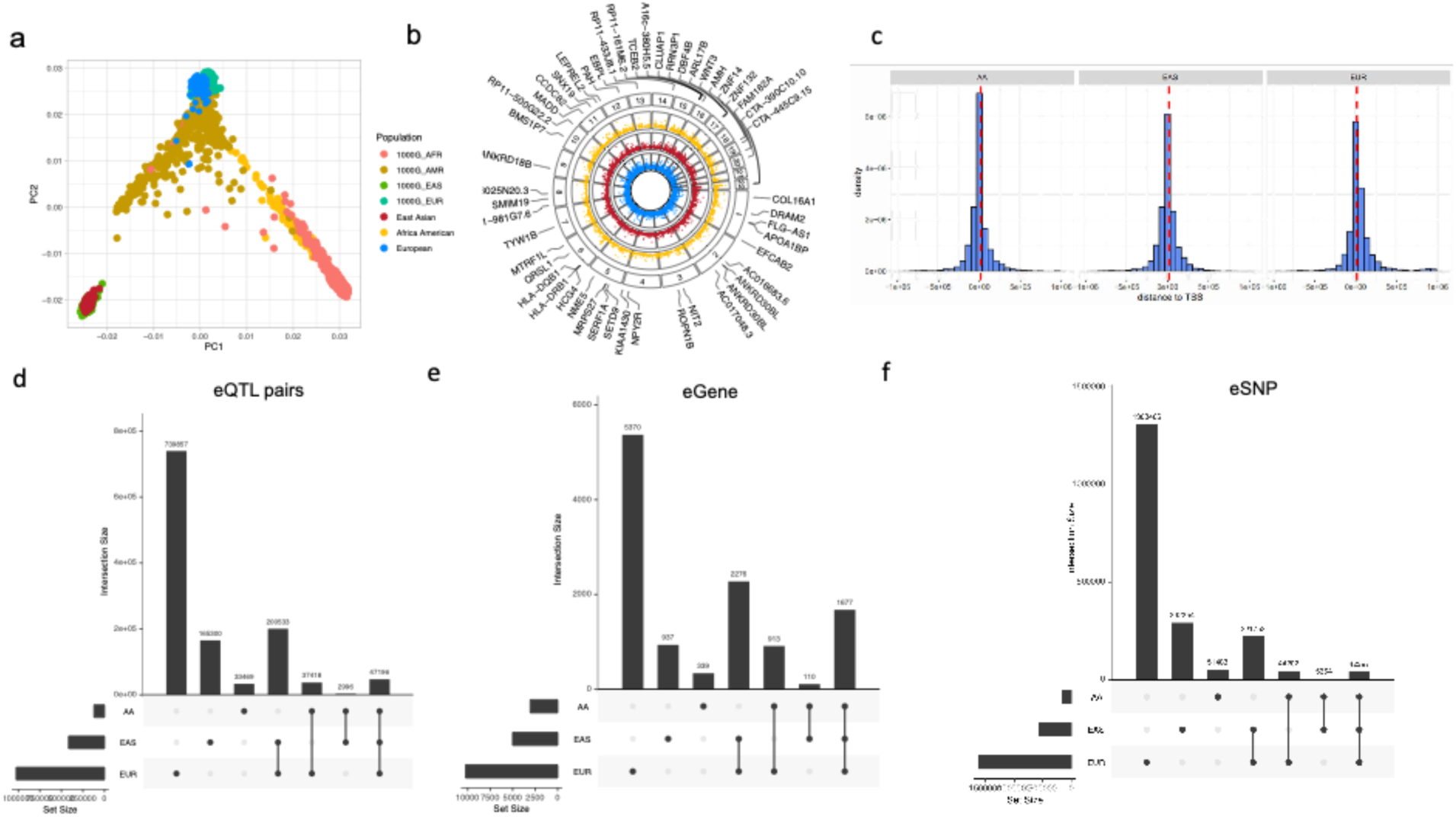
Identification and characterization of eQTLs. a) PCA plot showing the population structure of individuals in our study as well as the 1000 Genomes Project. AFR: African; AMR: American; EAS: East Asian; EUR: European; b) Circos manhattan plot of significant eQTL genes among the three populations with highlighted top 50 fine-mapped eGenes. d-f) Upset plot showed overlap among the significantly associated eQTL pairs (d) eGenes (e) as well as eSNPs (f) between populations.

### Characterizing the cis-acting eQTLs in European, East Asian, and African American population

We separately conducted cis-eQTLs mapping in the EUR, EAS, and AA samples using a 5% empirical gene-level false discovery rate (FDR) threshold. This yielded 1,966,209 significant eQTL signals covering 11,622 genes (eGenes) and 1,226,769 SNPs (eSNPs) across the populations (see Fig. 2b and Supplementary Table 2). Specifically, we identified 1,616,818 eQTL signals spanning 10,236 eGenes and 1,025,004 eSNPs for EUR; 562,058 eQTLs incorporating 5,000 eGenes and 416,025 eSNPs for EAS; and 143,736 eQTLs covering 3,039 eGenes and 121,079 eSNPs for AA. To identify credible SNP sets harboring plausible causal variants in cis-eQTLs, we applied a fine-mapping method named SuSiE^10^ to each population’s eQTL results. The results showed 966 credible SNP sets for 757 eGenes in the EUR cohort, 826 sets for 726 eGenes in EAS, and 847 sets for 746 eGenes in AA (Supplementary Tables 3-5).

To investigate the genomic features of these cis-eQTLs, we evaluated the SNP distributions and locations relative to various functional regions. 20% of cis-eQTLs in both EUR and non-EUR populations were located within 10kb of transcription start site (TSS) regions (Fig. 2c). According to the chromatin states predicted by Genomic Regulatory Elements and Gwas Overlap algoRithm (GREGOR)^11^ for prefrontal cortical tissue, eSNPs from the non-EUR populations were significantly enriched in TSSs, promoters, and transcribed regulatory promoters or enhancers (P_Bonferroni_ < 0.05), identical to the observation in the EUR data. Moreover, using transcription factor binding site (TFBS) annotation for 51 TFs, 46 and 49 TFs were significantly enriched with cis-eQTLs in the AA and EAS populations respectively (P_Bonferroni_ < 0.05). All these TFBS were also significantly enriched with cis-eQTLs in the EUR population.

To maximize the power of our population-based datasets, we employed METAL to amalgamate the cis-eQTLs data from all three populations. This meta-analysis generated 598,193 eQTLs, correlating 436,456 eSNPs with 5,209 eGenes (with a stringent P<2.5e-6) as depicted in Fig. 2b. However, meta-analysis does not consider the LD differences among the populations. We thus used SuSiEx^16^ to identify likely causal variants regulating expression by incorporating the LD reference data from different populations. In total, SuSiEx identified 2,121 credible SNP sets for 1,801 eGenes in the 3-population combined data. Further details from the meta-analysis and fine-mapping results can be found in Supplementary Table 6.

### Population shared eQTLs showed similar regulatory effect across population

To evaluate the effect sizes across populations, we conducted a correlation test of effect size values between the EUR and non-EUR populations. The effect sizes of the shared eQTLs between the non-EUR and EUR were highly concordant (0.910 for eQTLs comparing EAS to EUR and 0.944 for AA to EUR; Fig 3a). To assess the robustness of the concordant effect size, we examined eQTL slopes in the different populations. We looked at eQTLs obtained from the nominal, permutation, and conditional tests, separately. eQTLs with smaller p-values or larger effect sizes showed greater consistency across populations (Extended Data Fig. 3). Considering that sample size and heterogeneity may influence the results, we randomly down-sampled the EUR data to match the size of the non-EUR data. The results were similar to the results comparing all samples, showing highly concordant effect sizes across populations (see Fig. 3c, Extended Data Fig. 3). In addition, we compared the slope of the down-sampled EUR data with GTEx data (also of EUR). We randomly sub-sampled 100 times and obtained a distribution of correlation values (R^2^). The mean R^2^ was 0.94, which was not significantly different from the correlation between the EUR and non-EUR population. We therefore concluded that the effect sizes of eQTLs in diverse populations were mostly stable across human populations.

**Fig. 3:**
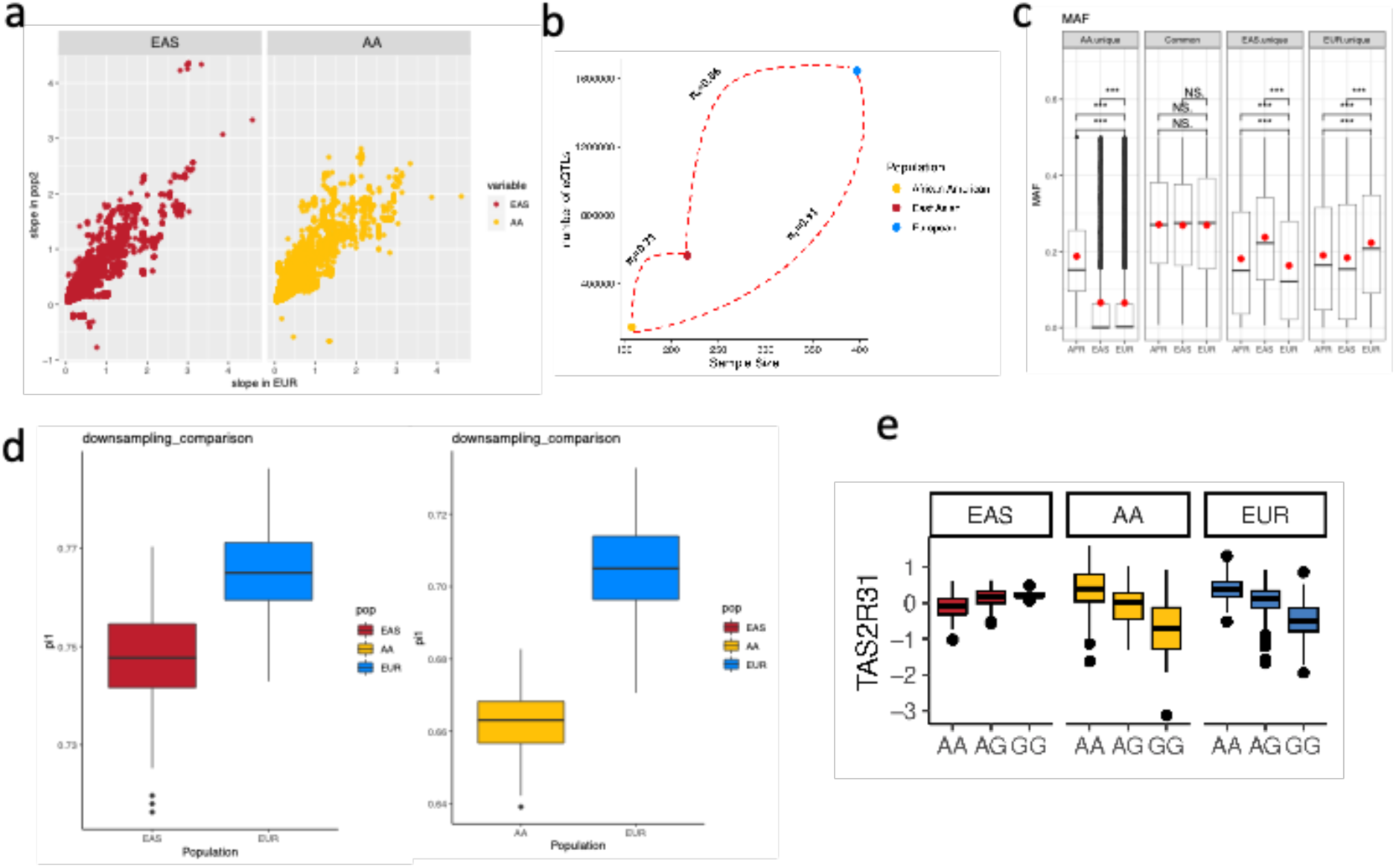
Analysis of the regulatory patterns.a) relationship between sample size and the # of detected eQTLs. b) Effect sizes for common (MAF> 1%) sentinel cis-eQTLs across EA and AA populations; c) Comparison of MAF between population-shared and non-EUR unique eSNPs; d) downsampling to estimate π_1_ between non-European and European eQTLs . e) example of opposite effect eQTL *TAS2R31*-rs2599400.

We evaluated the replicated rate (π_1_), which gauges the true positive rate for the eQTLs identified in the non-EUR populations that were also associated in the EUR population. The replicated rate was π_1_(EAS-EUR) = 0.86 and π_1_(AA-EUR) = 0.91 (see Fig.3b). The π_1_ for the non-EUR populations in EUR was slightly but significantly lower than the π_1_ between two EUR cohorts, as represented by the GTEx cis-eQTL data (prefrontal cortex) in our EUR eQTL data (π_1_(GTEx-EUR) = 0.86, p-value = 0.023). To ensure a fair comparison of the replication rate of detected cis-eQTLs in non-EUR data, we adjusted the EUR data to reflect the smaller sample size of the non-EUR data. This adjustment enabled us to determine how many non-EUR cis-eQTLs were confirmed in the adjusted EUR dataset. The adjusted results revealed a concordant trend: the replicated rate between different populations was still slightly lower than that within the same population assuming the same sample size (EUR-nonEUR average π_1_ = 0.68, EUR_adjusted_-EUR average π_1_ = 0.72, p-value = 0.037) (see Fig. 3d).

### Population differences in brain cis-eQTLs are mainly caused by differences in SNP allele frequency while differences in effect size are small and uncertain

Here we defined those eQTLs that were exclusively observed in a single population as population-specific eQTLs. Upon analyzing the cis-eQTLs overlapping between populations, we identified 343,737 cis-eQTLs that were exclusively observed in the non-EUR populations, as detailed in Supplementary Table 2. This number represents approximately 17% of all eQTL pairs. These eQTLs involved 1,276 genes (about 10% of all eGenes) and 198,769 SNPs (around 16% of all eSNPs, Fig. 2d-f.). Specifically, there were 292,254 cis-eQTLs involving 165,300 eSNPs and 937 eGenes that were observed only in the EAS population and 51,483 cis-eQTLs involving 33,469 eSNPs and 339 eGenes that were observed only in the AA population.

To further characterize these non-EUR-specific eQTLs, we analyzed the variance, taking into account both the eQTL slope (effect size) and differences in allele frequency between populations. We found that more than 90% of the population differences in variance were attributable to differences in allele frequency. Moreover, to delve deeper into the distinctive characteristics of the eQTLs exclusive to the non-EUR groups, we leveraged two statistics, the fixation index (FST) and the minor allele frequency (MAF), retrieved from the 1000 Genomes Selection Browser^12^. A high FST value indicates that the measured locus has diverged over time in the populations. As expected, eSNPs detected only in the EAS or AA population displayed a significantly elevated FST when juxtaposed against eSNPs shared across populations (Wilcoxon test P < 2.2e-16). Meanwhile, the non-EUR-specific eSNPs showed higher MAF values in their respective source populations (Wilcoxon test P < 2.2e-16;) than in EUR. Of the 343,737 eQTLs absent in the EUR data, 309,363 were likely due to inadequate statistical power because they have smaller MAF in the EUR than non-EUR population. Furthermore, 70,980 were rare in the EUR population (MAF_1KG_ < 0.05).

For the rest the eQTLs for which population differences could not be explained by differences in MAF, a test for differences in eQTL slopes (effect sizes) was also conducted between the EUR and non-EUR populations. The z-score of each independent eQTLs from conditional analysis was calculated based on effect size and its standard deviation. Here the null hypothesis was that the difference in eQTL effect size between the populations equals zero. No eQTL pairs detected by conditional analysis could reject the null hypothesis. We then investigated if any eQTLs exhibited opposite effect directions across populations. None of the independent eQTLs from the conditional analysis displayed such effects. We relaxed our eQTL threshold using a nominal p-value < 0.05. Only 534 eQTLs involving eighteen genes exhibited opposing eQTL effects between the EUR and non-EUR populations. For example, the bitter taste receptor gene *TAS2R31* showed opposite directions of eQTLs in EAS and EUR (Fig. 3e), which could be replicated using the blood eQTLs from a previous study^7,13^.

In conclusion, the variance in population differences can be largely attributed to differences in allele frequency. The influence of effect size differences, on the other hand, appears to be minimal and inconclusive.

### Brain eQTLs from matched population can improve interpretation of SCZ GWAS

To determine if eQTLs detected from a specific population could explain the disease GWAS signals and SNP-based disease heritability better than eQTLs from non-matching populations, we undertook a two-step analysis. Firstly, we gathered SCZ GWAS summary statistics for the EUR, EAS, and AA populations from previously published studies^14–16^. We employed the partitioned LD-score regression (LDSR)^17^ approach to assess the GWAS signal enrichment of these eQTLs. eQTLs identified in the EAS population demonstrated a markedly higher enrichment in EAS-based GWAS signals than eQTLs identified in the EUR population. Conversely, eQTLs identified in the EUR population showed a greater enrichment for EUR-based GWAS signals than the eQTLs from the EAS cohort. Both of these enrichments were statistically significant (Welch Modified Two-Sample t-Test P-value < 0.001, Fig. 4a and 4b).

**Fig. 4:**
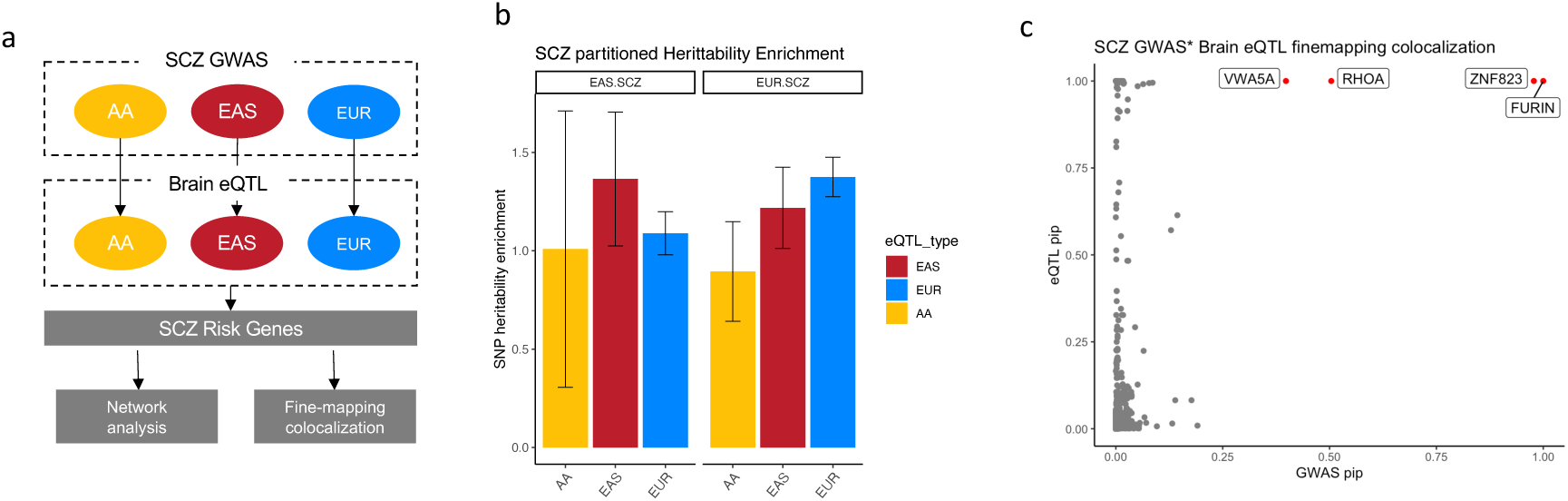
Explanation of SCZ GWAS signals and prioritization of candidate genes. a) integration strategies; b) GWAS enrichment results from LDSR. ***: Welch Modified Two-Sample t-Test P < 0.001; c) Fine-mapped colocalization results. Each point represents an eQTL pais, the x-axis represents the GWAS pip for that eSNP, and the y-axis represents the eQTL pip for that eSNP. Red points represent pip_GWAS_*pip_eQTL_>0.1 and are labeled with the eGene

Besides the SNP heritability enrichment of all eQTLs, we also compared the significance of the GWAS signals for population-unique eSNPs. We found that population-unique eSNPs tended to have smaller p-values of disease association (i.e., stronger associations) in the corresponding population than in the mismatched population (Welch Modified Two-Sample t-Test P < 0.001), indicating the ability of population-unique eSNPs to explain the disease association and propose the relevant gene, which will be missed when only one population is studied.

### SCZ risk genes identified using eQTLs and GWAS from non-EUR population

To uncover risk genes and pathways for SCZ in non-EUR populations, we used TWAS, regulatory Trait Concordance (RTC)^18^, and summary data-based mendelian randomization (SMR)^19^ to prioritize SCZ candidate risk genes in non-EUR populations and compared them with risk genes identified in the EUR (see Methods). In total, we prioritized ten risk genes in the EAS (Supplementary Tables 7-9). It is worth noting that our TWAS analysis of AA data did not reveal any significant associations. This lack of association in AA data might be attributed to the relatively small sample size available from the AA SCZ GWAS.

Seven SCZ candidate risk genes (*SFXN2*, *RP11-282018.3*, *CYP17A1*, *VPS37B*, *DENR*, *FTCDNL1*, and *NT5DC2*) uniquely discovered in the EAS population were assessed for allele frequency. From these seven genes, two of them were located in chromosome 10 and five of them were located in the chromosome 12. The eSNPs for these genes showed lower allele frequency in the EUR population than in EAS. For instance, the GWAS signal chr12:123286491:A:G in the gene *VPS37B* was found to be significant in the EAS population with a high AF of 0.48. In contrast, this association was not significant in the EUR population with a markedly lower allele frequency of 0.04. A parallel pattern emerged with the eSNP for *VPS37B*, with a markedly higher frequency (chr12:123306558:G:A, MAF=0.24) in EAS than in the EUR (MAF = 0.04). These results further confirm that allele frequency differences between populations can explain most of the discrepancies between the EUR and EAS GWAS and the eQTL results (Supplementary Table 10).

### Novel potential SCZ regulatory variations were refined utilizing brain eQTLs from non-EUR population

Three of the ten risk genes identified in the EAS population were shared with the EUR population (*CNNM2*, *C12orf65*, and *MPHOSPH9*), but differences in genetic architecture between populations were still apparent. For example, two distinct significant SNPs in EAS and in EUR were associated with SCZ on chromosome 10 (GWAS_EUR_: chr10:104850632:G:A with GWAS p-value = 6.4E-13; GWAS_EAS_: chr10:104657300:T:C with GWAS p-value = 5.4E-12). Using eQTLs with GWAS signals in EAS and EUR separately, colocalization and SMR analysis prioritized these two distinct GWAS SNPs to the same risk gene *CNNM2* in the two populations, respectively.

To further investigate if these signals are located within any regulatory elements, we utilized the non-coding variant annotation database (NCAD)^20^ to annotate their regulatory information. Our findings revealed that all the EAS GWAS risk SNPs are situated in enhancer regions (Supplementary Table 11). The EAS GWAS risk SNPs near the *CNNM2* showed the strongest evidence. Furthermore, we used the Lineage-specific Brain Open Chromatin Atlas^21^ to investigate whether this enhancer region shows different effects in major brain cell types. The results did not show any cell type differences, which indicates the enhancer effect exists universally in major brain cell types (Extended Data Fig. 4a-c). Integrating these insights, we discovered strong evidence for multiple regulatory regions among the EAS eSNPs-chr10:104654577:T:C, which have a high LD with *CNNM2* GWAS SNPs (LD R^2^ = 1, p-value < 0.00001). Additionally, our dual luciferase reporter assay results confirmed that the EAS eSNPs C-allele at chr10:104654577:T:C significantly enhances luciferase activity compared to the reference vector, as detailed in the Extended Data Fig. 4d-e.

### High-confidence putative causal variants of SCZ using multi-ancestry brain eQTLs

To identify high-confidence putative causal variants from multiple populations, we applied colocalization to our fine-mapped eQTLs and SCZ GWAS signals. In total, we identified four SNP-gene-disease triplets in which the SNP colocalized with both gene expression and SCZ GWAS (Supplementary Table 12, PiPcoloc = PiP_GWAS_ × PiP_cis-eQTLs_ > 0.1). The top genes with PIP_coloc_ > 0.1 include *FURIN*, *ZNF823*, *RHOA*, and *VWA5A* (Fig. 4c). As an example, we identified the strongest putative SCZ causal SNP for *FURIN-*chr15:91426560:G:A. This SNP is located in the 3’ untranslated regions (UTRs) of *FURIN*. Notably, this variant did not reach genome-wide significance in the EAS population (P = 1.06E-3) likely due to limited statistical power. Our result strongly supported that this causal variant is shared across populations, with causal probabilities of 1. Previous study has also implicated the variant in both the EUR and EAS populations^13^.

## Discussion

In this study, we have created a brain transcriptome resource and identified eQTLs in the prefrontal cortex, specifically focusing on non-European populations. Our findings address the initial inquiries raised in the introduction. Firstly, we investigated the driver behind the variation in brain eQTLs across different populations. We found that differences in allele frequency and LD are instrumental in connecting disease susceptibility to gene expression regulation. This finding greatly augments our comprehension of genetic influences on gene expression in the human brain. Secondly, when examining brain eQTLs from diverse populations, we gained power to explain the GWAS heritability, uncover novel risk genes, and fine-map risk variants. We observed a pronounced enrichment of disease heritability among eQTLs in matched populations. In the non-EUR cohort, the allele frequencies and LD configurations facilitated the identification of seven novel SCZ risk genes. Additionally, we identified four high-confidence putative causal SCZ variants. These results highlight the utility of studying non-European cohorts.

Population differences appear to be more pronounced at the allele frequency level but are less so at the effect size level. In general, the estimated π_1_ of eQTLs from non-EUR populations in EUR is lower compared to the rate observed between down-sampling-EUR and the EUR population cohort. Despite the relatively small sample size and statistical power, we still detected a significant number of 343,737 cis-eQTLs including 232,254 EAS eQTLs and 51,483 AA eQTLs that were significant only in the non-EUR populations. While over half of eSNPs in our non-EUR dataset were population-unique, 80% of eGenes identified in the non-EUR were also eGenes in the EUR data, but associated with different SNPs. The consistency of our observations with prior research involving diverse populations, including studies on gene expression^22,23^, methylation^24^, and chromatin accessibility^25^, confirms the shared regulatory patterns across different populations.

Interestingly, some eQTLs showed contrasting effects across populations. ∼0.1% of the non-EUR-specific eQTLs displayed opposing directions in effect size. A notable example of this is the eQTL rs2599400-*TAS2R31*, which showed opposite effects in different populations. Blood eQTLs from EAS^26^, EUR, and AFR^7^ also support this observation. Prior studies have underscored the population-specific variations in *TAS2R31*, linking these variations to differing sensitivity to the bitter taste^27^. It is important to further investigate these discrepancies in gene expression regulation among populations as they influence phenotypes. However, it should be noted that lack of replication across all these studies limited our power to detect opposite-effect eQTLs.

Our findings underscore that enhancing genetic ancestral diversity is more efficient for power gain than increasing the sample size within large-scale eQTLs datasets. Through our benchmarking of eQTLs across three populations, we have established robust capabilities for identifying eQTLs with a MAF greater than 0.2 and an effect size of 0.6 (Extended Data Fig. 5). Our power analysis indicates that more than 30,000 individuals of European ancestry is needed to uncover all eQTLs with MAF of 0.01 in this population, based on the estimated effect size of eQTLs exclusively observed in non-EUR populations (Extended Data Fig. 5). For example, one the eQTL pair (chr12:123306558:G:A-VPS37B) would require 3,215 EUR samples based on the power estimate because of the low frequency in the EUR population (MAF = 0.04). However, the MAF of this eSNP is 0.22 in EAS, which reduces the required sample size from 3,215 to 246. Thus, incorporating a more diverse population would not only reveal numerous regulatory variants that are rarer in EUR but more prevalent in non-EUR groups. Advancing towards a broader, more diverse human reference dataset will facilitate more comprehensive investigations into the impact of human demography on eQTL detection, thereby deepening our understanding of the distribution and influence of genetic regulation in the human brain.

Differences in the genetic architecture underlying gene expression can help us to prioritize new risk genes. Notably, prior research has reported that disease-associated loci tend to be skewed towards variants with higher allele frequency in the discovery population, indicating that limited statistical power may result in “missing” disease-association signals. Incorporating diverse samples can enhance our ability to uncover the etiology of the disease. In our study, we identified seven new SCZ risk genes using the non-EUR population, including *VPS37B* in the EAS population. *VPS37B* is associated with calcium-dependent protein binding, providing new evidence to support the involvement of the calcium-related pathway in SCZ risk in the EAS population^28^. Another interesting candidate highlighted in our study was *CYP17A1* (RTC = 0.99). The corresponding GWAS signal was significant in the EAS and EUR populations (P_EAS_ = 4.5E-8; MAF_EAS_ = 0.48; P_EUR_ = 2.6E-13; MAF_EUR_ =0.30), while the corresponding eSNP in EAS population (MAF=0.48) showed extremely low frequency in EUR population (MAF<0.001). *CYP17A1* notably serves as an enzyme important for the production of glucocorticoids and sex hormones, such as estrogen, which have been linked to schizophrenia^29–31^ .

Besides enhancing the power for detecting risk genes, the inclusion of brain eQTLs from diverse populations improves the ability to fine-map SCZ GWAS loci, identifying new regulatory variants which have the potential to regulate downstream gene expression. This approach aids in interpretation, thereby facilitating subsequent computational and experimental functional investigations. Our result revealed a potential novel regulatory region near the population-shared risk gene *CNNM2*. This discovery showcases the power of leveraging diverse populations.

By leveraging the multi-ancestry information, trans-ancestry fine-mapping also helped us identify high-confidence putative causal variants. In addition to previously validated genes, our study uncovered another significant finding at chr3:50297330:A:G-*RHOA*-SCZ through trans-ancestry colocalization. *RHOA* encodes a member of the Rho family of small GTPases, pivotal in signal transduction cascades by toggling between inactive GDP-bound and active GTP-bound states.

Some limitations of our study merit attention. Our sample size is still relatively small. Our analysis suggests that expanding the sample size would capture a larger set of eQTLs. The small sample size could have significantly impacted the comprehensiveness of our findings. The modest sample size of the AA eQTL dataset and the SCZ GWAS cohort likely led to the failure of TWAS in the AA population.

In conclusion, we presented a novel genome-wide map of human brain gene expression regulation. Importantly, this resource bridges the gap between neuropsychiatric GWAS and brain gene expression profiling in non-European populations. Our study emphasizes the significance of this new atlas of brain gene expression regulation in non-European populations for advancing our understanding of human diversity, addressing health disparities, and developing precision medicine.

## Materials and methods

### Sample collection and sequencing

We collected 217 prefrontal cortical samples of Han Chinese ancestry from the National Human Brain Bank for Development and Function^32,33^; the samples were handled according to the standardized operational protocol of the China Human Brain Banking Consortium, under the approval of the Institutional Review Board of the Institute of Basic Medical Sciences, Chinese Academy of Medical Sciences, Beijing, China (Approval Number: 009-2014).

We sequenced 217 samples following the BGISEQ-500 protocol outsourced to BGI. 1μg genomic DNA was randomly fragmented by Covaris, the fragmented DNA was selected by Agencourt AMPure XP-Medium kit to an average size of 200-400bp, followed by adapter ligation, PCR amplification, and the products were recovered by the AxyPrep Mag PCR clean up kit. The double-stranded PCR products were heat-denatured and circularized by the splint oligo sequence. The single-strand circle DNA (ssCir DNA) was formatted as the final library and qualified by QC. Sequencing was performed on BGISEQ-500 platform with an average depth of 10X.

Total RNA was extracted from the brain tissue using Trizol (Invitrogen, Carlsbad, CA, USA) according to manufacturers’ instructions. Then, total RNA was qualified and quantified using a Nano Drop and Agilent 2100 bioanalyzer (Thermo Fisher Scientific, MA, USA). Ribo-zero method was used to remove the rRNA. Purified mRNA was fragmented into small pieces with fragment buffer at an appropriate temperature. The cDNAs were purified by magnetic beads. After purification, A-Tailing Mix and RNA Index Adapters were added by incubating to carry out end repair. The cDNA fragments with adapters were amplified by PCR, and the products were purified by Ampure XP Beads. The library was validated on the Agilent Technologies 2100 bioanalyzer for quality control. The final library was amplified with phi29 (Thermo Fisher Scientific, MA, USA) to make DNA nanoball (DNB), DNBs were loaded into the patterned nanoarray and single end 50 base reads were generated on BGISEQ500 platform.

### Data quality control

Raw sequencing reads were filtered to get clean reads by using SOAPnuke (v1.5.6)^34^, and FastQC^35^ was used to evaluate the quality of sequencing data via several metrics, including sequence quality per base, sequence duplication levels, and quality score distribution for each sample. The average quality score for overall DNA and RNA sequences was above 30, indicating that a high percentage of the sequences had high quality.

### Variant identification

Clean DNA sequencing reads were mapped to the human reference genome hg19 (GRCh37) using BWA-MEM algorithm (BWA v. 0.7.128)^36^. Ambiguously mapped reads (MAPQ <10) and duplicated reads were removed using SAMtools v. 1.29^37^ and PicardTools v. 1.1 respectively. Genomic variants were called following the Genome Analysis Toolkit software (GATK v. 3.4.4.6) best practices.

### Population validation, imputation, and filtering

We used PLINK to infer the genomic ancestry of each sample in this study by combining our genotype data and the genotype data from the 1000 Genomes Project^38^; no sample was excluded. Using Michigan Imputation Server^41^, EAS genotypes were imputed into the 1000 Genomes Project phase 3 EAS reference panel by chromosome and subsequently merged. Imputed genotypes were filtered for LD R^2^ < 0.3, Hardy-Weinberg equilibrium p-value < 10e-6 and MAF < 0.05, resulting in ∼ 6 million autosomal SNPs.

For AA population, genotypes were imputed into the 1000 Genomes Project phase 3 AA reference panel by chromosome and subsequently merged. To further confirm the ancestry of the African American samples, all AA samples were evaluated for their ancestry with three broad population groups with PC1 ≥ 25% AFR and < 25% AMR, < 25% EAS, < 25% SAS; clustering of individuals in each broad population group with the 1000 Genomes Project reference populations are shown in Fig.2a.

### Sex check and sample swap identification

The sex of each sample was inferred with SNPs using PLINK. In the EAS cohort, two samples were identified as sex-mismatched and were subsequently removed in downstream analysis. Quality control was performed on genotypes using sample Binary Alignment Map (BAM) files to detect any sample identity swaps between the RNA and DNA experiments. The QTLtools match function^39^ confirmed that all samples were appropriately matched.

### Gene expression quantification and quality control

The RNA-sequencing reads were mapped using STAR (2.4.2a)^40^ and the genes and transcripts quantification was performed using RSEM (1.3.0)^41^. Raw read counts were log-transformed using R package VOOM^42^, thereafter filtering those with log2(CPM) < 0 in more than 75% of the samples. Mitochondrial DNA and X and Y chromosome-derived transcripts were excluded. Samples with a Z-score (measured for inter-sample connectivity) less than -3 were also discarded. Finally, quantile normalization was utilized to equalize distributions across samples.

### Covariate selection

To measure technical covariates, quality control metrics were collected using STAR, PicardTools v1.139 and RNASeQC. Principal components of the metrics data were calculated and included as SeqPCs for covariate selection. Hidden covariates were measured using probabilistic estimation of expression residuals (PEER)^43^ and found to be significantly correlated with technical and biological covariates such as experimental batch, RNA Integrity Number (RIN), sex, and age of death. Based on the Bayesian information criterion (BIC) score, redundant covariates were removed to avoid overfitting. A forward and backward selection procedure was followed, and PEER hidden factors and known covariates were added. The covariate with the higher BIC score was selected for subsequent QTL mapping. PEER was run with varying numbers of inferred hidden factors: 5, 10, 15, 20, 25, 30, 35, 40, 45 and 50, independently.

### cis-eQTL mapping

Cis-eQTL mapping was performed using QTLtools, accounting for PEER factors, with a defined cis window spanning one megabase upstream and downstream of the gene/intron cluster body. Using the PEER from gene expression matrix, twenty hidden covariates were identified and adjusted. To detect all available QTLs, QTLtools was conducted in nominal pass mode. To identify the best nominal associated SNP per phenotype, QTLtools was executed in the permutation pass mode. Additionally, to identify SNPs with independent effects on regulating gene expression, QTLtools was run in the conditional pass mode. SNPs with P-values below the threshold of the permutation threshold is classified as significant QTLs.

To address potential sample size disparities that could impact the results, the EUR data were randomly sampled with various sample size (150, 200, 250, 300, 350, and 400) and applied the same analytical pipeline while exploring the relationship between sample size and number of QTLs.

### eQTL meta-analysis

The SNP-gene pairs with a genome-wide P value reaching the threshold of 10e-6 for each population were collected and performed the meta-analysis. Standard fixed-effects meta-analysis were used to combine all data into a single regression model by METAL^44^. The meta-analysis assumes a fixed-effects size, as well as constant error variance, across all data.

### eQTL fine-mapping

The initial step of fine-mapping involved using the in-sample LD of the three populations. We extracted common variants with MAF > 5% from each group and used PLINK to determine the LD regions of these common variants for each population. To eliminate strand flipping and alignment issues, multi-allelic variations and indels were removed. Next, SuSiEx was applied to merge the eQTLs summary statistics from the three groups. Credible set is defined as a set of putative causal variants. A credible set was discarded if it lacked genetic variants reaching genome-wide significance (p < 1e-6) in either the population-unique eQTLs or cross-population meta-eQTLs. By considering prior knowledge and the observed data, this method provides a posterior probability (PIP) for each variant being the causal one in the associated region. Variants with high PIPs are then considered strong candidates for functional follow-up studies.

### Functional enrichment

To determine functional enrichment, GREGOR^48^ was performed to test the eQTL enrichment. GREGOR calculated the enrichment value based on the observed and expected overlap within each annotation. To conduct our analysis, the 15-state ChromHMM model BED (Browser Extensible Data) files from the Roadmap Epigenetics Project^45^, and 78 consensus transcription factor and DNA-protein binding site BED files existing in multiple cells were downloaded. Fifty binding proteins showed cortical brain expression in EAS and AA populations data^46^.

### The fraction of shared eQTLs between non-EUR and EUR populations

Sharing rate was assessed based on significant eQTLs in the discovery dataset by estimating the proportion of true associations (π_1_) on the distribution of corresponding p-values of the overlapping eQTLs in the replication dataset^47^.

### F_ST_ and MAF analysis

Fixation index (F_ST_) was estimated using vcftools following the Weir and Cockerham approach for each eSNP^48^. The population-divergent SNPs were defined as those with F_ST_ >= 0.05 and population-shared SNPs as those with F_ST_ < 0.05. To generate the list of population-unique QTLs and population-shared QTLs, we collected the overlap of eQTLs from the pairwise comparisons of the list of AA eQTLs, EAS eQTLs, and EUR eQTLs. Finally, Fisher’s exact test was performed between population-unique QTLs and population-shared QTLs to test the contribution of MAF in the QTL comparison.

### Variance explained

Variance explained, which combines the effect size (beta) and frequency of the allele (f), can be considered an approximate measure of a causal variant’s importance within a population. Variance is approximated using the formula 2f(1 – f)log(beta)²/(π²/3)^49^. Although these variants often exhibit similar odds ratios across populations, their allele frequencies may differ. By considering both the effect size (OR) and the frequency of the risk allele (f), the variance explained offers a valuable approximation of a causal variant’s significance within a given population.

### Power estimation

We used R to calculate the sample size needed to achieve a given power level in a chi-square test, based on an assumed effect size and a significance threshold. This analysis starts by setting initial values for power, effect size, and p-value threshold. Then, the critical chi-square statistic required to meet the power level is computed. A function, *calculate_ncp*, is defined to calculate the non-centrality parameter from the p-value and degrees of freedom, adjusting for the critical chi-square statistic. Subsequently, the non-centrality parameter is computed for the given power and p-value threshold. Another function, *af_n_relation*, is created to determine the relationship between allele frequency and sample size, incorporating the effect size and the non-centrality parameter. Finally, the code iteratively solves for the sample size corresponding to a range of allele frequencies, thus enabling the determination of the necessary sample size for different allele frequencies to maintain the specified power level in the chi-square test.

### Partitioned LDSR

Partitioned LD score regression v1.0.1^50^ was used to measure the enrichment of GWAS summary statistics in each functional category by accounting for LD. Brain QTL annotations were created by eSNP, mapped to the corresponding 1000 Genome reference panel. LD scores were calculated for each SNP in the QTL annotation using an LD window of 1cM in 1000 Genomes European Phase 3 and 1000 Genomes Asian Phase 3 separately. Enrichment for each annotation was calculated by the proportion of heritability explained by each annotation divided by the proportion of SNPs in the genome falling in that annotation category. We then applied Welch Modified Two-Sample t-Test on enrichment values generated from QTLs in the two populations.

### Colocalization

Conditional association was used to test for evidence of colocalization. This method compares the p-value of association for the lead SNP of an eQTL before and after conditioning on the GWAS hit. The equation for the regulatory trait concordance (RTC) Score is as follows: RTC= (N_SNPs_ in an LD block/Rank_GWAS_SNP_)/ N_SNPs_ in an LD block. The rank denoted the number of SNPs, which when used to correct the expression data, has a higher impact on the QTL than the GWAS SNPs. RTC values close to 1.0 indicated causal regulatory effects. A threshold of 0.9 was used to select causal regulatory elements.

Colocalization of fine-mapped variations from complex traits and cis-eQTL correlations were performed. Based on complex trait and cis-eQTL fine-mapping data, a posterior inclusion probability of colocalization for a variant was calculated as a product of PIP for GWAS and PIP for the cis-eQTLs (PIPcoloc = PIP_GWAS_ * PIP_cis-eQTLs_).

### Summary-data-based Mendelian randomization

SMR^19^ was applied on SCZ GWAS summary data to prioritize candidate genes. Significant QTLs identified in the previous analysis (FDR < 0.05), were combined with filtered GWAS summary data (p < 5e-8) to perform the SMR test. In general, we used the default parameters suggested by the developers of the SMR software. These included the application of heterogeneity independent instruments (HEIDI) testing, filtering out hits that arose from significant linkage with pleiotropically associated variants (LD cutoff of P = 0.05 in the HEIDI test, as suggested by SMR). Genes with an empirical P that passed Bonferroni correction in the SMR test and a P > 0.05 in the HEIDI test were considered as risk genes.

### Prioritizing genes underlying GWAS hits using TWAS

In this research, we initially developed gene expression prediction models for distinct populations using MetaXcan software^51^. Following this, we integrated these models with GWAS (Genome-Wide Association Studies) summary statistics specifically focused on schizophrenia. This integration aimed to generate gene-level z-scores representing the association of the genetically-determined expression for a gene from its prediction model with the phenotype. TWAS enabled us to compute P-values and subsequently prioritize genes in relation to their association with schizophrenia risk.

### Plasmid construction

We obtained the 55 bp SNP-centered DNA sequence from UCSC Genome Browser (GRCh38/hg38), then added the sticky end of restriction enzymes KpnI and NheI at both ends of the 55bp sequence to synthesis primers. Primer annealing to obtained double-strand sequence and then inserted into pGL3-Promoter Vector (Promega) using FastDigest enzymes (ThermoFisher) and T4 DNA Ligase (Invitrogen). We valid the vector sequence using sanger sequence by primer GLP2 and RVP3.

### Dual Luciferase Reporter Assay

For transfection, we used SH-SY5Y and HS-683 cells to perform the experiments. Transfecting cells at 50-60% confluency, cells were co-transfected with 500ng reconstruction vector and 10ng pRL-TK using Lipofectamine 3000 Transfection Reagent ( ThermoFisher ) in 24 well plates. After 48h transfection, using Dual Luciferase Reporter Assay Kit (Promega) to measure the firefly luciferase activity and renilla luciferase activity, the luminescence was detected using Tube Luminometer (Berthold Sirius).

## Data availability

The raw sequence data reported in this paper have been deposited in the Genome Sequence Archive in BIG Data Center, Beijing Institute of Genomics (BIG), Chinese Academy of Sciences, under accession numbers HRA000108, HRA000108 that can be accessed at https://bigd.big.ac.cn/gsa-human. eQTLs summary results can be downloaded while requesting to the corresponding author.

## Code availability

Codes are available at https://github.com/liusihan/population-compare-pipeline.

## Acknowledgments

We thank all donors and their families. Tissue was provided by the Human Brain Bank, Chinese Academy of Medical Sciences & Peking Union Medical College, Beijing, China, the Chinese Brain Bank Center, and the Xiangya School of Medicine Brain Bank. We acknowledge Stanley Center for Psychiatric Research for supporting Hailiang Huang and Yu Chen working on this project.

## Author contributions

Y.C. and S.L. wrote the manuscript, analyzed the data and performed all computations. F.W. and Y. J. contributed to eQTLs analysis of EUR population. Z.R., F.D., C.H and M.L. extracted DNA and RNA, as well as collected sample information. R.D., Y.X., and R.K. substantively revised the manuscript. Z.N., S.X. and H.H. participated in the design of comparing the brain regulatory architecture. K.Y., W.Q., C.M., X.Y., A.B., J.D., J.H., B.T. and C.L. provided primary sequenced samples and data including their clinical information. C.M., C.L. and C.C. conceived, designed and supervised the study and modified the manuscript.

## Ethnics

Ethics Committee of Central South University gave ethical approval for this work (2015031007).

---

**Extended Data Figure 1:**
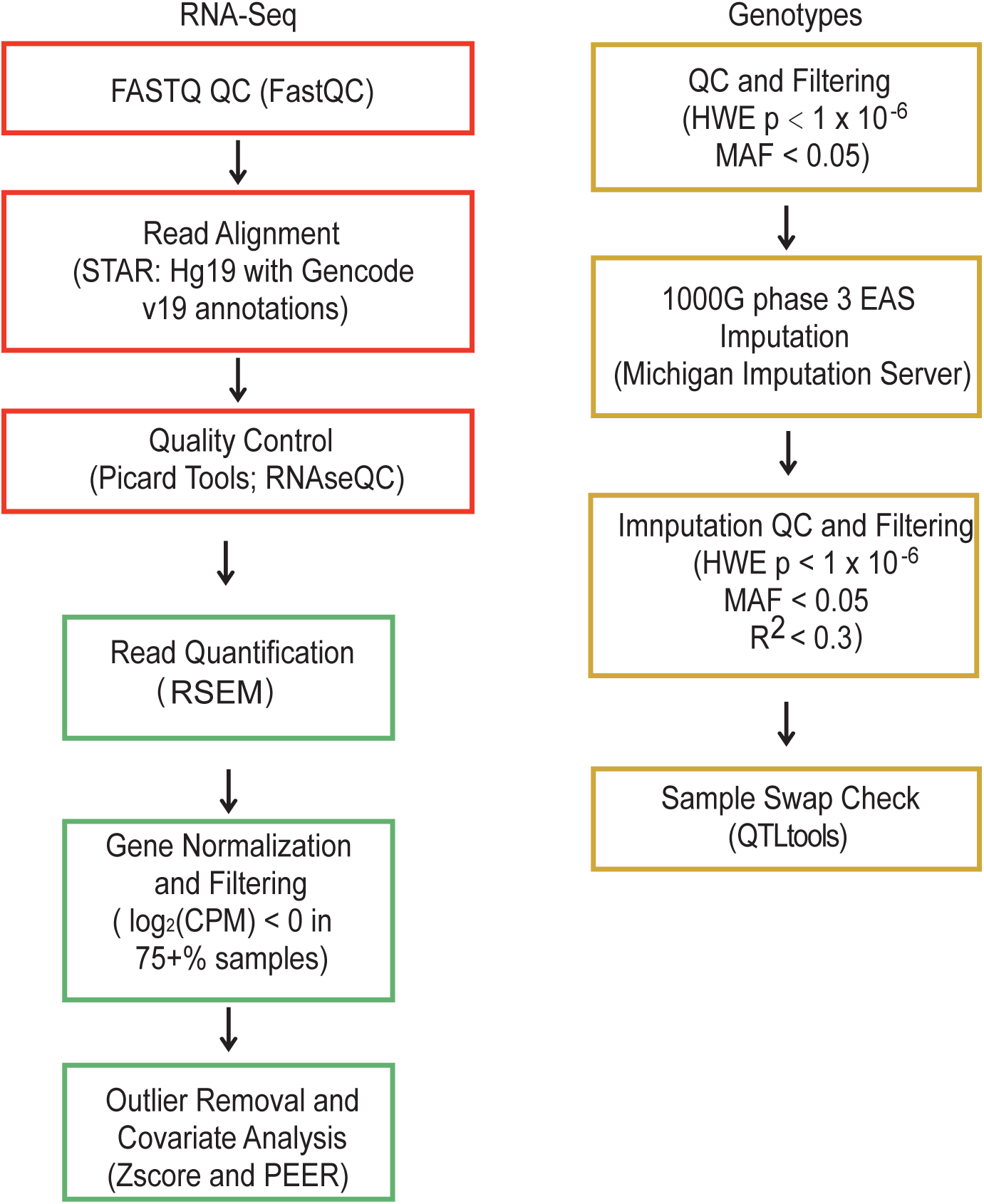
Overview of methods and QC pipeline for EAS samples.

**Extended Data Figure 2:**
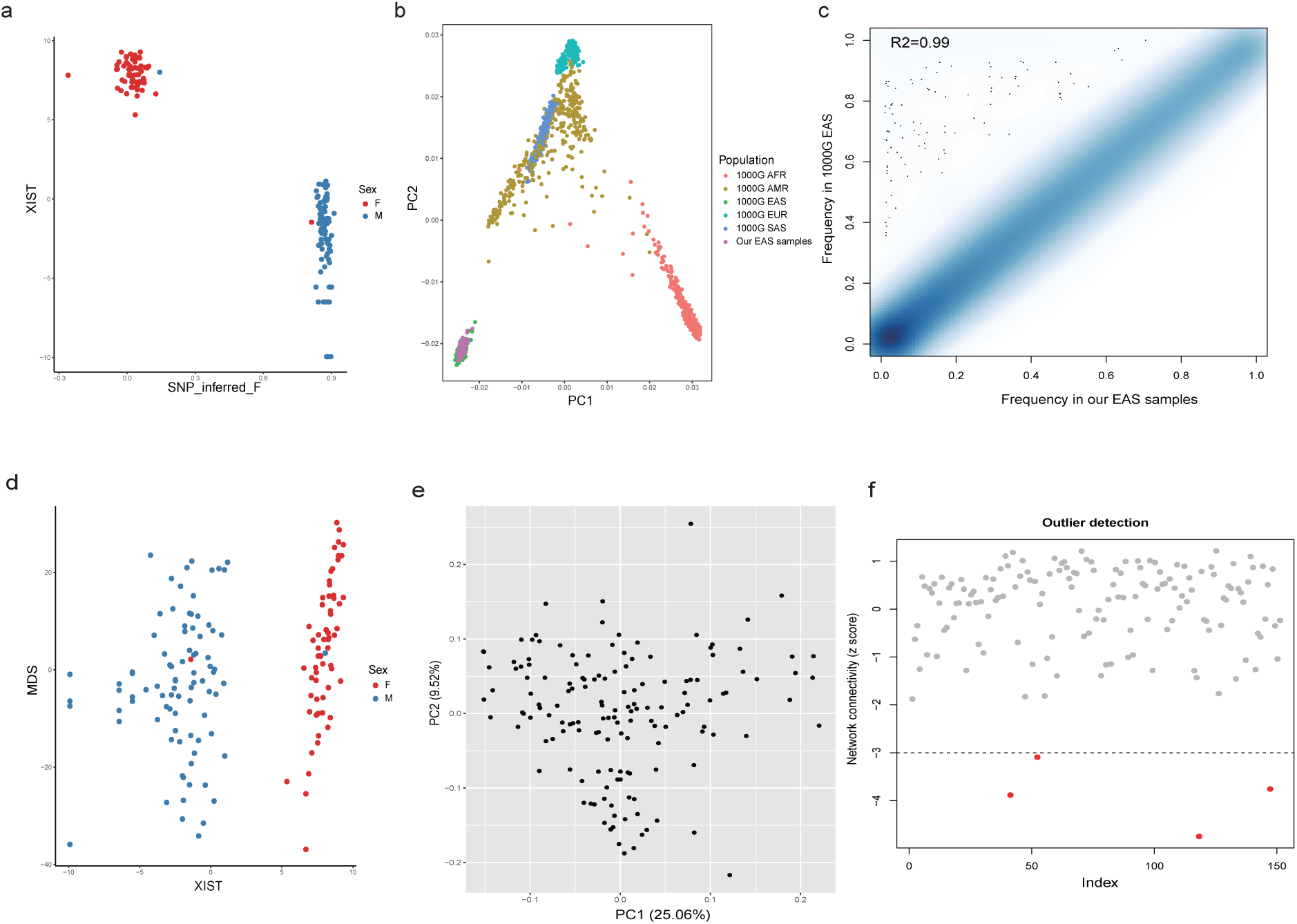
Preprocessing of RNA-sequencing and whole-genome sequencing (WGS) data of EAS samples. a, Sex-mismatch checked by WGS data. b, Population PCA plot with 1000G genotype data. c, Imputation accuracy. d, Sex-mismatch checked by Xist expression. e, PCA plot for EAS samples. f, Distribution of Z-score.

**Extended Data Figure 3:**
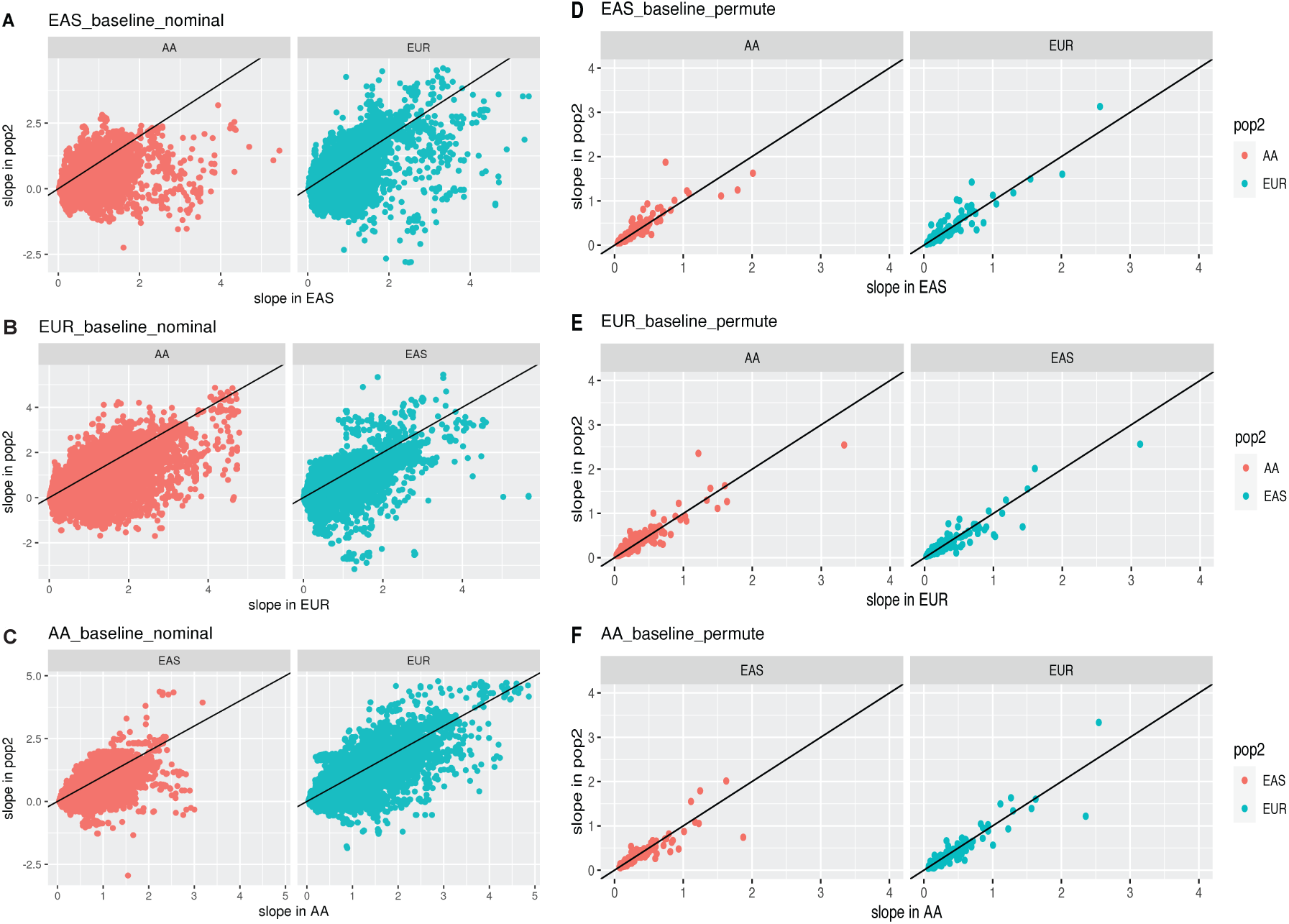
Effect size correlation of population-shared eQTLs between EUR and non-EUR population. (a-c) nominal pass; (d-f) permutation pass

**Extended Data Figure 4:**
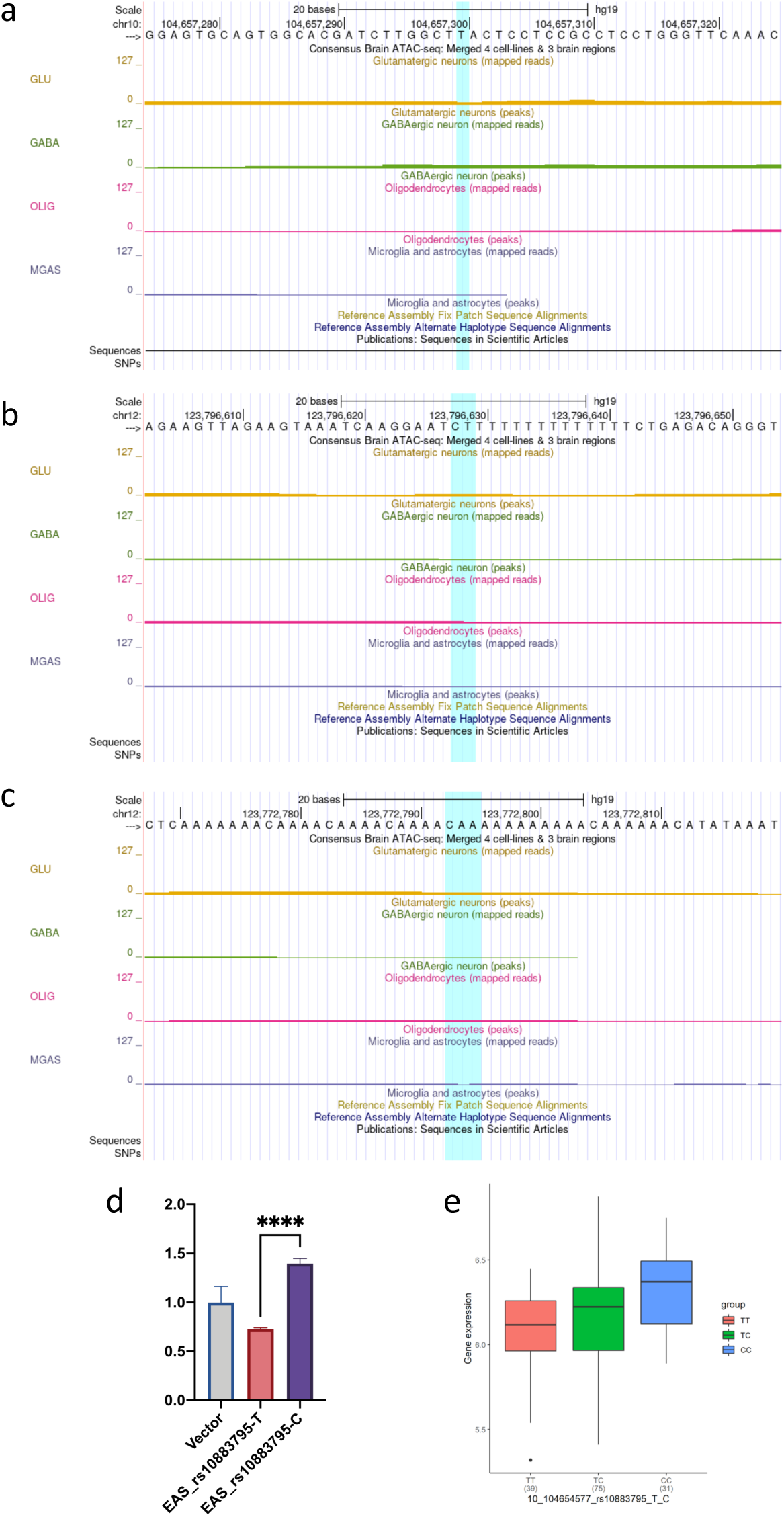
Regulatory effect across glutamate neuron, GABA neuron, oligodendrocytes, and microglia for new regulatory SNPs within population-shared risk genes for (a) CNNM2, (b)C12orf65, (c) MPHOSPH9. (d) Dual luciferase reporter assay for EAS eSNP at risk gene CNNM2. (f) eQTL result for the eSNP and expression risk gene CNNM2 in EAS cohort

**Extended Data Figure 5:**
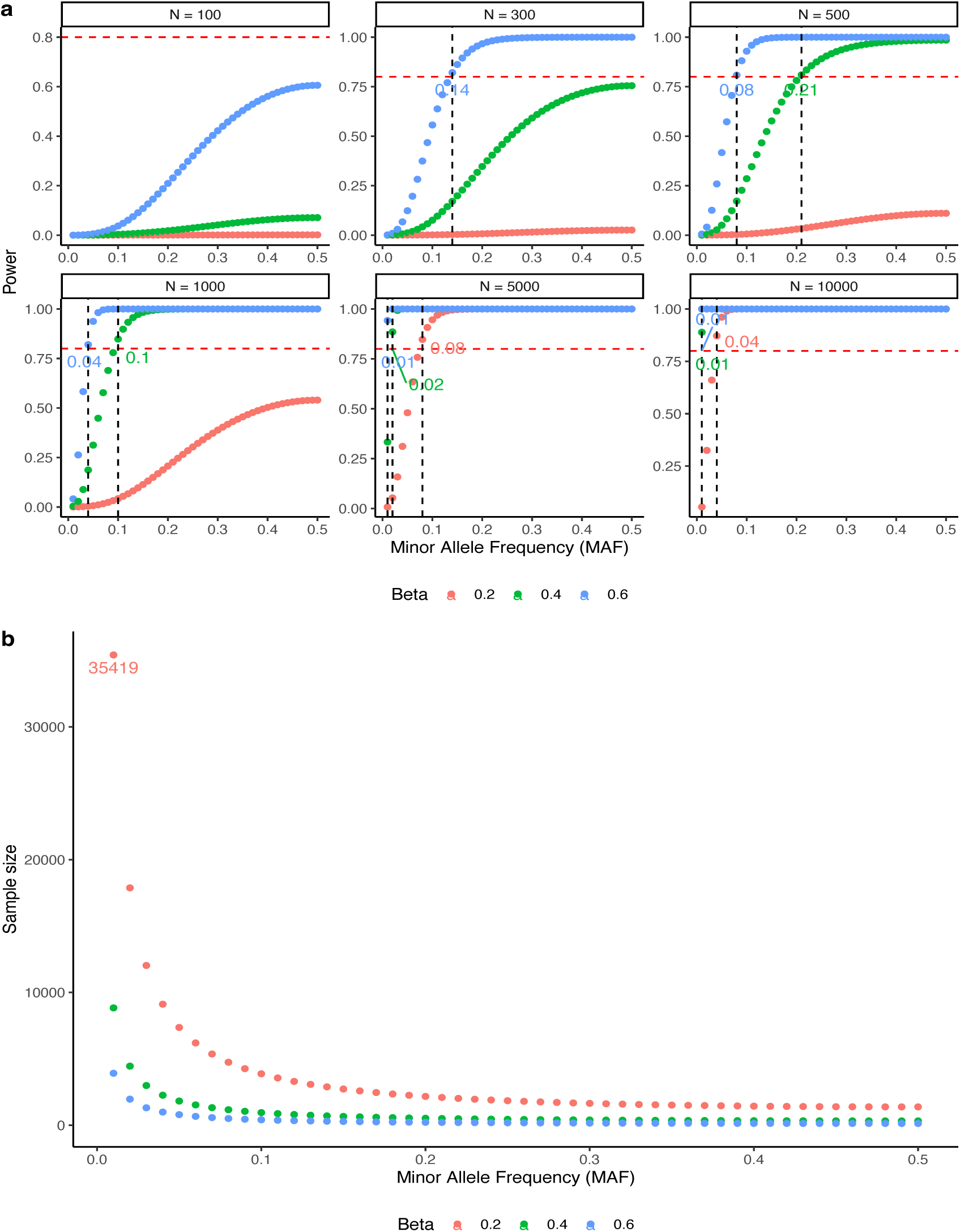
The sample size required for well-powered brain eQTL detection in diverse populations. (a) The percentage of brain eQTLs detected power under different sample sizes and effect sizes is shown as a function of log-scaled sample size. (b) The required sample size achieving 80% power based on the effect size estimated form non-EUR specific eQTLs.

